# Potentially effective drugs for the treatment of COVID-19 or MIS-C in children: a systematic review

**DOI:** 10.1101/2021.07.20.21260827

**Authors:** Zijun Wang, Siya Zhao, Yuyi Tang, Zhili Wang, Qianling Shi, Xiangyang Dang, Lidan Gan, Shuai Peng, Weiguo Li, Qi Zhou, Qinyuan Li, Joy James Mafiana, Rafael González Cortés, Zhengxiu Luo, Enmei Liu, Yaolong Chen

**Affiliations:** Evidence-based Medicine Center, School of Basic Medical Sciences, Lanzhou University, Lanzhou, China; Lanzhou University Institute of Health Data Science, Lanzhou, China; School of Public Health, Lanzhou University, Lanzhou, China; Department of Respiratory Medicine, Children’s Hospital of Chongqing Medical University, Chongqing, China; National Clinical Research Center for Child Health and Disorders, Ministry of Education Key Laboratory of Child Development and Disorders, China International Science and Technology Cooperation Base of Child Development and Critical Disorders, Children’s Hospital of Chongqing Medical University, Chongqing, China; Chongqing Key Laboratory of Pediatrics, Chongqing, China; The First School of Clinical Medicine, Lanzhou University, Lanzhou, China; Paediatric Intensive Care Unit, Hospital General Universitario Gregorio Marañón, Calle Doctor Castelo 47, 28007 Madrid, Spain; WHO Collaborating Centre for Guideline Implementation and Knowledge Translation, Lanzhou, China; Lanzhou University GRADE Center

**Keywords:** children, COVID-19, MIS-C, glucocorticoids, intravenous immunoglobulin, remdesivir

## Abstract

**Introduction:** The purpose of this systematic review is to evaluate the efficacy and safety of using potential drugs: remdesivir and glucocorticoid in treating children and adolescents with COVID-19 and intravenous immunoglobulin (IVIG) in treating MIS-C.

**Methods:** We searched seven databases, three preprint platform, ClinicalTrials.gov, and Google from December 1, 2019, to August 5, 2021, to collect evidence of remdesivir, glucocorticoid, and IVIG which were used in children and adolescents with COVID-19 or MIS-C.

**Results:** A total of six cohort studies and one case series study were included in this systematic review. In terms of remdesivir, the meta-analysis of single-arm cohort studies have shown that, after the treatment, 37.1% (95%CI, 0.0% to 74.5%) experienced adverse events, 5.9% (95%CI, 1.5% to 10.2%) died, 37.2% (95%CI, 0% to 76.0%) needed extracorporeal membrane oxygenation or invasive mechanical ventilation. As for glucocorticoids, the results of the meta-analysis showed that the fixed-effect summary odds ratio for the association with mortality was 2.79 (95%CI, 0.13 to 60.87), and the mechanical ventilation rate was 3.12 (95%CI, 0.80 to 12.08) for glucocorticoids compared with the control group. In terms of IVIG, the two included cohort studies showed that for MIS-C patients with more severe clinical symptoms, IVIG combined with methylprednisolone could achieve better clinical efficacy than IVIG alone.

**Conclusions:** Overall, the current evidence in the included studies is insignificant and of low quality. It is recommended to conduct high-quality randomized controlled trials of remdesivir, glucocorticoids, and IVIG in children and adolescents with COVID-19 or MIS-C to provide substantial evidence for the development of guidelines.

## 1. Introduction

It is over a year and a half since the outbreak of coronavirus disease 2019 (COVID-19), and during this period, studies on COVID-19 are continuously emerging [1,2]. Researchers have paid much attention to drug therapy all the time [3]. Recent studies on COVID-19 drugs and clinical guidelines have focused primarily on adult patients but less attention on children and adolescents. Although children and adolescents with COVID-19 seem less susceptible and have milder symptoms once infected, they are also at risks of advancing to severe stages [4]. Children and adults are known to have physiological differences [5]; thus, many effective COVID-19 drugs for adults may not suitable for children. Among these drugs, remdesivir, glucocorticoids, and intravenous immunoglobulin (IVIG) in children and adolescents have been controversial.

Remdesivir is a broad-spectrum antiviral medication that can integrate into the RNA strand of severe acute respiratory syndrome coronavirus-2 (SARS-CoV-2) and prematurely terminate the ribonucleic acid (RNA) replication process [6]. The World Health Organization (WHO) living guideline for COVID-19 [7] and the United States guideline for pediatric COVID-19 [8] have contradicting recommendations for the treatment of children and adolescents, based on evidence from randomized controlled trials of adults. At the same time, the status of original studies of remdesivir in children and adolescents with COVID-19 is unclear.

Glucocorticoids are the most widely used and effective anti-inflammatory and immunosuppressive agents in clinical practice. They have the potential to reduce the severity of lung inflammation in patients with severe COVID-19 [9,10]. Glucocorticoids are affordable, easy to administer, and readily available globally [11]. The WHO living guidance on glucocorticoids for COVID-19 [12] recommends systemic glucocorticoids to treat adult patients with severe COVID-19. However, the living guidance further suggests that the recommendation is underrepresented in children and adolescents with COVID-19.

IVIG is a recommended first-line therapy for Kawasaki disease because it produces anti-inflammatory effect, which reduces coronary artery abnormalities and myocarditis in patients with Kawasaki disease [13]. MIS-C (multi-system inflammatory syndrome in children) is a newly defined clinical syndrome associated with SARS-CoV-2 infection characterized by fever, systemic inflammation, and multiple organ dysfunction. Several case definitions of this novel inflammatory condition have been published by the WHO [14], the US Center for Disease Control and Prevention (CDC) [15], and the United Kingdom of Great Britain Royal College of Pediatrics and Child Health (RCPCH) [16]. The clinical features of MIS-C are similar to those of Kawasaki disease, toxic shock syndrome, sepsis, and macrophage activation syndrome [17]. Hence, the application of IVIG in the treatment of MIS-C is a potential drug choice [18], but the evidence of the application of IVIG in MIS-C treatment is still unclear.

Therefore, we aimed to determine the efficacy and safety of using: 1. remdesivir in treating children and adolescents with COVID-19, 2. glucocorticoids in treating children and adolescents with severe COVID-19, 3. IVIG in treating children and adolescents with MIS-C. Furthermore, provide evidence to support the development of clinical practice guidelines.

## 2. Methods

Six researchers in three groups of two (Group 1: Zijun Wang, Qianling Shi; Group 2: Siya Zhao, Qi Zhou; Group 3: Yuyi Tang, Weiguo Li) retrieved and selected studies, extracted and analyzed data, and interpreted the results. Group 1 focused on remdesivir in treating children and adolescents with COVID-19, Group 2 focused on glucocorticoids in treating children and adolescents with severe COVID-19 and Group 3 focused on IVIG in treating children and adolescents with MIS-C. We reported our study in accordance to the Preferred Reporting Items for Systematic Reviews and Meta Analyses (PRISMA) 2020 guidelines. [19] (Supplementary File 1)

### 2.1. Search strategy

Two researchers in each group independently searched for literature using MEDLINE (via PubMed), Web of Science, the Cochrane library, China Biology Medicine (CBM), China National Knowledge Infrastructure (CNKI), Wanfang Data, and WHO COVID-19 database (https://search.bvsalud.org/global-literature-on-novel-coronavirus-2019-ncov/), ClinicalTrials.gov (https://clinicaltrials.gov/), MedRxiv (https://www.medrxiv.org/), BioRxiv (https://www.biorxiv.org/), SSRN (https://www.ssrn.com/index.cfm/en/), and Google. The electronic search was supplemented by manually examining the reference lists of the identified studies. In addition, emails were sent to the authors of studies to request available data that may be useful for our systematic review. The data search was from December 2019 to August 2021 without language limitations.

The researchers in groups 1, 2, and 3 used “remdesivir,” “corticosteroids,” and “intravenous immunoglobulin,” and its derivatives as retrieval terms, respectively. The terms were also combined with “COVID-19” and its derivatives using “AND”. For question 3, “MIS-C” and its derivatives were added as retrieval terms and combined with “AND” to improve the accuracy of the search. The search strategy can be found in Supplementary File 2.

### 2.2. Eligibility criteria

#### 2.2.1. Inclusion criteria

Clinical question 1 (remdesivir)

Randomized controlled trials, non-randomized controlled trials, cohort studies, and case series of children and adolescents (≤ 18-year-old) with COVID-19 treated with remdesivir.

Clinical question 2 (glucocorticoids)

Randomized controlled trials, non-randomized controlled trials, cohort studies, and case series of COVID-19 children and adolescents (≤ 18 year) patients treated with glucocorticoids.

Clinical question 3 (IVIG)

1. The study population must meet the diagnostic criteria for MIS-C, and the included patients were not restricted by age, gender, disease course, race, region, and other factors.
2. The interventions/exposure included IVIG (intravenous immunoglobulin) vs. placebo or other treatment, or IVIG combined with other treatment vs. basic treatment.
3. Inclusion of studies was not restricted by the type of publication.

#### 2.2.2. Exclusion criteria

Clinical question 1 (remdesivir)

1. Studies that failed to show the efficacy of remdesivir.
2. Case series that remdesivir was not administered to all the patients or subgroup comparison of remdesivir was unavailable.
3. Full text not available (example, studies inaccessible for download, conference abstract).
4. Duplications.

Clinical question 2 (glucocorticoids)

1. Studies that failed to show the efficacy of glucocorticoids.
2. Case series that glucocorticoid was not administered to all the patients or subgroup comparison of glucocorticoid was unavailable.
3. Full text not available (example, studies inaccessible for download, conference abstract).
4. Duplications.

Clinical question 3 (IVIG)

1. In vitro studies (example, animal experiments, in vitro experiments).
2. Full text not available (e.g., studies inaccessible for download, conference abstract).
3. Duplications.

### 2.3. Study selection

Two researchers in each group independently screened literature using the EndNote citation management software, and any disagreements were resolved by discussion. Before the formal screening process, researchers in each group randomly selected 50 studies to undertake a pilot study selection and ensure consistency in understanding the inclusion and exclusion criteria. Researchers used the inclusions and exclusions criteria first to screen the studies’ title and abstracts and excluded irrelevant literature. Then, the full text of the literature was reviewed to include the final eligible studies. Finally, the reasons for exclusion were recorded. The details of study selection are shown in the PRISMA 2020 flow diagram (Supplementary File 3).

### 2.4. Data extraction

Two researchers in each group extracted data independently in pairs, using a predefined data extraction form. Disagreements regarding the data extraction were resolved by discussion. The following information was extracted from the included studies: 1) baseline characteristics: author, year of publication, country, journal, number of included patients, gender, age, study design, and medication taken for COVID-19; 2) data extracted for clinical question 1: adverse events, severe adverse events, mortality, extracorporeal membrane oxygenation (ECMO) or invasive mechanical ventilation (IMV), length of hospital stay, hospital discharge, and symptom duration; 3) data extracted for clinical question 2: mortality, mechanical ventilation, and duration of pediatric intensive care unit (PICU) admission; and 4) data extracted for clinical question 3: number of patients who had treatment failure or secondary acute left ventricular dysfunction, number of patients who needed second-line treatment or hemodynamic support, the duration of PICU stay, isovolumic relaxation time, and the time to recovery of left ventricle ejection.

For dichotomous variables, the data of the number of events and the total of events were extracted. For continuous variables, mean, standard deviation, and the number of included patients were extracted. The median, quartile, maximum values, and minimum values were converted into mean and standard deviation using methods of estimating math [20]. Studies were excluded from the meta-analysis if the primary data was unavailable and showed the results of descriptive analysis of those studies.

### 2.5. Risk of bias assessment

Two reviewers in each group independently assessed the risk of bias of all included studies, and discrepancies were resolved by consensus. The risk of bias of the included randomized controlled trials was assessed using Cochrane’s risk of bias tool [21]. Potential sources of bias are examined according to six domains (including seven items): selection bias, performance bias, detection bias, attrition bias, reporting bias, and other biases. Each item was assessed as “low risk of bias,” “high risk of bias,” or “unclear.” The risk of bias of included non-randomized controlled trials was assessed using the tool of ROBINS-I [22], which contains seven items (confounding, selection of participants into the study, classification of the intervention, deviations from intended interventions, missing data, measurements of outcomes, and selections of the reported result), each of which was assessed as “low risk,” “moderate risk,” “serious risk,” “critical risk,” and “no information”. The Newcastle-Ottawa quality assessment scale [23,24] was used to assess the risk of bias of cohort studies. The scale contains eight items in three domains: selection, comparability, and outcome. The items were rated with an asterisk. The Quality Appraisal Checklist for Case Series Studies developed by the Institute of Health Economics was used to assess the risk of bias of case series studies [25]. The checklist contains twenty items in eight domains: study objective, study population, intervention and co-intervention, outcome measure, statistical analysis, results and conclusions, competing interests and sources of support, and supplement. Each item was evaluated with “yes” or “no”.

### 2.6. Data synthesis

A meta-analysis using the STATA14 software when the outcomes of included studies were highly consistent and descriptive analyses when there was high heterogeneity of outcomes between the included studies. According to Cochrane Handbook, when the meta-analysis was conducted, a random-effects meta-analysis for all outcomes was presented [26]. For an included study with intervention group and control group, the odds ratios (ORs) and their 95% confidence interval (CI) were used to describe the effect of dichotomous variables while weighted mean differences (WMD) and their 95% CI were used to describe the effect of continuous variables. However, for an included study with only an intervention group, the effect sizes (ES) and their 95% CI were used to describe the effect of dichotomous variables while mean differences (MD) and their 95% CI were used to describe the effect of continuous variables. Statistical significance was set at <0.05 on both sides [27]. We used the chi-squared test and I^2^ statistic were used to assess the level of statistical heterogeneity between the included studies, with *p*<0.05 and I^2^ of less than 50% representing heterogeneity [27]. When substantial heterogeneity was detected, subgroup analyses by participant and study characteristics were used to compare pooled association estimates and heterogeneity. Furthermore, sensitivity analysis was used to detect potential outliers by omitting one estimate at a time and recalculating the pooled estimates. Publication bias was assessed through the funnel chart when the studies included in the meta-analyses were more than five [27].

### 2.7. Quality of the evidence assessment

Two reviewers in each group independently assessed the quality of evidence using the grading of recommendations assessment, development, and evaluation (GRADE) approach for meta-analysis. We created a “Summary of findings” table using GRADEpro to show effect estimates derived from the body of evidence (quality of evidence) by outcome [28,29]. Under the GRADE system, randomized controlled trials (RCTs) were initially assessed as high quality and observational studies as low quality. However, they were downgraded for reasons such as the risk of bias, inconsistency, indirectness, imprecision, publication bias, or upgraded for reasons such as the large magnitude of effect, dose-response gradient, and plausible confounding [30-35]. Thus, the quality of studies was rated as “high,” “medium,” “low,” and “very low,” reflecting the extent to which we are confident in the effect estimates.

Due to the peculiarity and public health significance of COVID-19, this study was not registered on the international registration platform PROSPERO.

## 3. Results

### 3.1. Study selection and characteristics

For clinical question 1, a total of 7292 records were retrieved from the databases and other methods. A total of three cohort studies were included, two of them was included from the database and one of them were unpublished studies obtained by data request [36-38]. For clinical question 2, 8025 records were retrieved. A cohort [39] and case series [40] study was included by reading the title, abstract, and full text. For clinical question 3, 3657 records were retrieved, and four cohort studies [41-44] were finally included. The detailed screening process for each clinical question is shown in Supplementary File 3.

### 3.2. Study characteristics

A total of 626 patients from the United States, Spain, France, and China were included in this study, of which the studies on IVIG were all from France (Table 1).

**Table 1.** Basic characteristics of the included studies

| Study | Region | Date | Study design | Total sample size | Sex (F/M) | Intervention |  | Control |  | Age, median (IQR), y | Drug |
| --- | --- | --- | --- | --- | --- | --- | --- | --- | --- | --- | --- |
|  |  |  |  |  |  | Sample size | Details | Sample size | Details |  |  |
| Méndez-Echevarría et al. 2020 [36] | Spain | Nov 16, 2020 | Single-arm Cohort | 8 | 3/5 | 8 | Children who weighed 40 Kg or more at screening received a single 200 mg dose on day one, following by a daily 100-mg dose from day 2 up to 10 days. For the rest of the children, a single dose of 5 mg/kg on day one was prescribed, followed by a daily dose of 2.5 mg/kg from day 2 up to 10 days. | NA | NA | 5 (0.3-11) | Remdesivir |
| **Munoz et al.**<br>**2021 [37]** | America | Mar 6,<br>2021 | Single-arm<br>Cohort | 27 | 15/12 | 27 | ≥40 kg received RDV 200 mg IV<br>Loading dose followed by RDV<br>100mg IV daily for up to total of 10<br>day of treatment; <40 kg received<br>RDV 5 mg/kg loading dose<br>followed by RDV 2.5 mg/kg IV<br>daily for up to total of 10 day of<br>treatment | NA | NA | 10 (0.2-17)<sup>#</sup> | Remdesivir |
| **Goldman et al.**<br>**2021 [38]** | America | July 10,<br>2020 | Single-arm<br>Cohort | 77 | 31/46 | 77 | ≥40 kg received RDV 200 mg IV<br>Loading dose followed by RDV<br>100mg IV daily for up to total of 10<br>day of treatment; <40 kg received<br>RDV 5 mg/kg loading dose<br>followed by RDV 2.5 mg/kg IV<br>daily for up to total of 10 day of<br>treatment | NA | NA | 14 (0-17)<sup>#</sup> | Remdesivir |
| **García-Salido et al. 2020<sup>8</sup> [39]** | Spain | Nov 26, 2020 | Cohort | 61 | 23/38 | 40 | Glucocorticoids (Not specified) | 21 | No glucocorticoids | 7.5 (4.9)<sup>□</sup> | Glucocorticoids |
| **Sun et al. 2020 [40]** | China | Mar 19, 2020 | Case series | 8 | 2/6 | 5 | Glucocorticoids (Not specified) | 3 | No glucocorticoids | 6.8 (6.5)<sup>□</sup> | Glucocorticoids |
| **Ouldali 2021\* [41]** | France | Mar 2, 2021 | Cohort | 96 | 99/101 | 64 | IVIG (2 g/kg) alone as first-line therapy | 32 | IVIG (2 g/kg) and methylprednisolone [0.8 to 1 mg/kg every 12 hours (maximum of 30 mg for 12 hours) for 5 days or a bolus of 15 to 30 mg/kg/d of methylprednisolone for 3 days.] | 8.6 (4.7-12.1) | IVIG |
| **Belhadjer 2020 [42]** | France | Dec 8, 2020 | Cohort | 40 | NR | 18 | IVIG (2g/kg) alone as first-line therapy | 22 | a combination of IVIG (2g/kg once) and intravenous methylprednisolone (0.8 mg/kg/d for 5 days) | 8.6 (6.7-11.2) | IVIG |
| **Son 2021 [43]** | America | Jun 16, 2021 | Cohort | 206 | NA | 103 | IVIG 2 g/kg (1.7, 2) | 103 | IVIG 2 g/kg (1.7, 2); Methylprednisolone 2 mg/kg/day (1.5, 2.67) or Dexamethasone 0.3 mg/kg/day (0.15, 2) or Prednisolone 2 mg/kg/day (1, 2.1) | NA | IVIG |
| **McArdle 2021 [44]** | United Kingdom | Jun 16, 2021 | Cohort | 420 | NR | 173 | IVIG (Not specified) | 177<br>70 | IVIG+ glucocorticoids (Not specified)<br>glucocorticoids (Not specified) | NA | IVIG |

1 \* After propensity score matching; # Mean (Range); § Data of diagnosed patients from authors; □ Mean (Standard Deviation); NA: Not Applicable; NR: Not Report

### 3.3. Risk of bias assessment

The results of risk of bias are shown in Supplementary File 4. The GRADE quality summary of findings for all outcomes is shown in Supplementary File 5.

### 3.4. Outcome of analysis

#### 3.4.1. Remdesivir

One hundred and twelve patients in 3 single-arm cohort studies [36-38] reported the efficacy and safety of remdesivir in treating children and adolescents with COVID-19. The results from a published study showed (n=8) [36] that 75% of the patients were admitted to the PICU, 62.5% were on mechanical ventilation, and 12.5% were on noninvasive ventilation or high-flow oxygen. In another study (n=77) [38], all the patients were diagnosed with severe COVID-19, among which 50.6% were treated with mechanical ventilation and 26.0% with noninvasive ventilation or high-flow oxygen. In another study (n=27) [37], 22% of patients received mechanical ventilation and 26% received noninvasive ventilation or high-flow oxygen. The results from an unpublished study showed that 79% (61/77) of the patients had an underlying disease. The meta-analysis of 104 children and adolescents with COVID-19 who received remdesivir showed that 12.4% (95%CI, 6.1% to 18.8%, very low quality evidence) experienced obesity, 11.4% (95%CI, 3.5 to 19.4%, very low quality evidence) experienced asthma, 6.9% (95%CI, 0.0% to 19.4%, very low quality evidence) experienced immunosuppression/immunologic diseases, 13.3% (95%CI, 6.8% to 19.8%, very low quality evidence) experienced epilepsy, 2.8% (95%CI, 0.0% to 6.0%, very low-quality evidence) experienced sickle cell disease. The result of the meta-analysis showed that, after the treatment, 37.1% (95%CI, 0.0% to 74.5%, very low-quality evidence) experienced adverse events, like acute kidney injury (19%, 5/27), constipation (15%, 4/27), increased alanine transaminase (ALT) (11%, 3/27), Hyperglycemia (11%, 3/27), Hypertension (11%, 3/27), Pyrexia (11%, 3/27) [37] and Anemia (3%, 2/77) [38]. There were 16.2% (95%CI, 1.8% to 30.5%, very low-quality evidence) of them experienced serious adverse events, 5.9% (95%CI, 1.5% to 10.2%) died, 37.2% (95%CI, 0% to 76.0%, very low-quality evidence) needed ECMO or IMV.

#### 3.4.2. Glucocorticoids

A retrospective cohort and case series studies [39,40] comprising of 69 children or adolescents (age 7.41±5.08) with severe COVID-19 treated with glucocorticoids were included. There was no statistically significant association between glucocorticoids therapy and mortality (OR= 2.79, 95% CI, 0.13 to 60.87, very low-quality evidence), mechanical ventilation rate (OR = 3.12, 95% CI, 0.80 to 12.08, very low-quality evidence) or the duration of PICU admission (WMD = 2.0, 95% CI, -0.95 to 4.95, very low-quality evidence).

#### 3.4.3. IVIG

One cohort study [41] showed that 64 patients who received IVIG alone as first-line therapy had a treatment success rate of 62% (treatment failure defined as the persistence of fever two days after introducing first-line therapy or recrudescence of fever within seven days after the first-line therapy). Patients with more severe initial clinical presentation (initial acute left ventricular dysfunction, initial PICU care, and hemodynamic support requirement) received a combination of IVIG and methylprednisolone as first-line therapy. The result showed that IVIG combined with methylprednisolone could decrease the treatment failure (OR=0.25, 95%CI, 0.09 to 0.70, low-quality evidence), second-line treatment (OR=0.19, 95%CI, 0.06 to 0.61, low-quality evidence), hemodynamic support (OR=0.21, 95%CI, 0.06 to 0.76, low-quality evidence), the occurrence of secondary acute left ventricular dysfunction (OR=0.20, 95%CI, 0.06 to 0.66, low-quality evidence), and duration of PICU stay (4 vs. 6, *p*=0.005).

Another cohort study [42] included 22 MIS-C patients who received a combination of IVIG (2 g/kg) and methylprednisolone (0.8 mg/kg/d for 5d). They had a shorter recovery time from left ventricle ejection fraction (2.9d vs 5.4 d, *p*=0.002), isovolumic relaxation time (6.4d vs 20.6d, *p*<0.0001), and duration of PICU stay (3.4d vs 5.3d, *p*<0.05), in comparison with the 18 patients that received only IVIG (2 g/kg) as first-line therapy (Very low quality evidence).

Similarly, another cohort study [43] with larger sample size showed that IVIG plus glucocorticoids was associated with a lower risk of the composite outcome of cardiovascular dysfunction on or after day 2 than IVIG alone (17% vs. 31%; RR=0.56, 95%CI, 0.34 to 0.94, very low quality evidence). The risks of the components of the composite outcome were also lower: left ventricular dysfunction (RR=0.46, 95% CI, 0.19 to 1.15, very low quality evidence), shock resulting in vasopressor use (RR=0.54, 95% CI, 0.29 to 1.00, very low quality evidence), and the use of adjunctive therapy (RR=0.49, 95% CI, 0.36 to 0.65, very low quality evidence).

However, in the other study [44] with 456 patients who met the WHO criteria for MIS-C, the authors compared IVIG plus glucocorticoids(n=186) and glucocorticoids alone(n=78) with IVIG alone(n=246), and found modest evidence of benefit with glucocorticoids alone over IVIG. The primary outcomes were the receipt of inotropic support or mechanical ventilation on day 2 or later or death (IVIG plus glucocorticoids vs. IVIG: OR=0.95, 95%CI: 0.37 to 2.45; glucocorticoids vs. IVIG: OR=0.30, 95% CI: 0.10 to 0.85), and the reduction in the score for disease severity on the ordinal scale by day 2 (IVIG plus glucocorticoids vs. IVIG: OR=1.09, 95% CI: 0.53 to 2.23; glucocorticoids vs. IVIG: OR=1.95, 95% CI: 0.83 to 4.60).

## 4. Discussion

### 4.1. Key findings

A total of six cohort studies and one case series study were included in this systematic review. In terms of remdesivir, there was no controlled study to prove its efficacy and safety in treating children and adolescents with COVID-19. Single-arm cohort studies have shown that the incidence of adverse reactions, mortality, and mechanical ventilation rate in patients treated with remdesivir are relatively low. As for glucocorticoids, the meta-analysis results showed no statistically significant difference in the improvement of mortality and mechanical ventilation rate between the intervention and control group. In terms of IVIG, the two included cohort studies showed that for MIS-C patients with more severe clinical symptoms, IVIG combined with methylprednisolone could achieve better clinical efficacy than IVIG alone.

The use of remdesivir in COVID-19 patients is a controversial topic for both adults and children. A systematic review and network meta-analysis of adult patients based on randomized controlled trials showed that patients treated with remdesivir for 5 days had a higher rate of clinical improvement compared with placebo [OR = 1.68 (95% CI 1.18–12.40)]. The rate of discharge [10-day remdesivir versus control: OR = 1.32 (95% CI 1.09–1.60); 5-day remdesivir versus control: OR = 1.73 (95% CI 1.28–2.35)] and recovery [10-day remdesivir versus control: OR = 1.29 (95% CI 1.03–1.60); 5-day remdesivir versus control: OR = 1.80 (95% CI 1.31–2.48)] of patients treated for 5 and 10 days were higher than placebo. Nevertheless, there was no significant improvement in mortality [45]. Other systematic reviews of adult patients have reached similar conclusions [46]. Based on this, on October 22, 2020, the U.S. Food and Drug Administration (FDA) approved Veklury (remdesivir) for the treatment of COVID-19 in children and adolescents aged at least 12 years and weighing at least 40 kg requiring hospitalization [47]. It also approved an emergency use authorization of remdesivir to treat suspected or laboratory-confirmed COVID-19 in hospitalized pediatric patients weighing at least 3.5 kg but being either aged less than 12 years or weighing less than 40 kg [48]. The results of this systematic review showed that most of the children and adolescents included in this study had severe or underlying diseases, and the adverse events, mechanical ventilation rate, and mortality of the patients after treatment with remdesivir were low, but there was a lack of control group; thus, the quality of evidence was low. The search in ClinicalTrials.gov showed that few studies focused only on children or adolescents with COVID-19 treated with remdesivir [49].

The effectiveness of glucocorticoids in the treatment of adult patients with COVID-19 has been confirmed. The Randomized Evaluation of COVID-19 Therapy (RECOVERY) Collaborative Group published an RCT on The New England Journal of Medicine, and the results of the study showed that among patients hospitalized with COVID-19, the use of dexamethasone resulted in lower 28-days mortality [9]. The WHO Rapid Evidence Appraisal for COVID-19 Therapies (REACT) working group published a systematic review based on seven RCTs. Results showed that systemic glucocorticoids administered to critically ill COVID-19 patients were associated with 28-days lower mortality than usual care or placebo [50]. Based on the systematic review evidence, the WHO developed a living guideline on glucocorticoids to recommend systemic glucocorticoids in treating patients with severe COVID-19 [12]. The recommendation was intended for the average patient population. However, the evidence that supported the recommendation was unclear for the under-represented population, such as children in the considered trials, which supported the meta-analysis of the systematic review. The search in ClinicalTrials.gov showed that no registered clinical trials have included or specifically targeted children or adolescents except for the RECOVERY trial. Most children with COVID-19 have only mild symptoms [5,51], so it may be challenging to recruit critically ill children or adolescents to participate in clinical trials. The two studies included in this systematic review were observational studies with a small sample [39,40] which found that glucocorticoids could not reduce the death rate in children or adolescents with critical COVID-19. Nevertheless, high-quality randomized controlled trials are recommended to confirm the effectiveness of glucocorticoids in the treatment of critically ill children or adolescents with COVID-19.

MIS-C is a unique complication in children and adolescents with COVID-19, which has similar characteristics to those of Kawasaki disease, but based on the limited evidence, the immunopathology of MIS-C remains a challenge [52]. Admittedly, IVIG generally produces anti-inflammatory effects, mitigates coronary artery abnormalities, and serves as first-line therapy of Kawasaki disease [53]. Several MIS-C guidelines are published, and the treatment therapy is based chiefly on Kawasaki disease, where IVIG is recommended empirically as the first-line treatment [49-51]. Besides, IVIG combined with glucocorticoids is also suggested as adjuvant therapy for severe patients or intensive therapy for patients with refractory diseases [54]. Three cohort studies included in this study showed that IVIG combined with glucocorticoids had better efficacy in MIS-C treatment than IVIG alone. Two of the three studies indicated that patients in the IVIG plus glucocorticoids group had more severe symptoms such as acute left ventricular dysfunction, admission to PICU care, mechanical ventilation, etc. The result is in agreement with the guideline recommendation of the use of IVIG in children and adolescents with COVID-19. Current guidelines also indicate a lack of high-quality studies comparing IVIG with glucocorticoids in MIS-C [54-57]. Different from the aforementioned three studies, the other study included a glucocorticoid-only group and the results provided modest evidence of benefit with glucocorticoids alone over IVIG alone. However, when expanding the range of patients to MIS-C and also those with any suspected inflammatory illness after SARS-CoV-2 infection, the data showed no differences between treatment with glucocorticoids or IVIG as single agents or between the single-agent and dual-agent treatments. The different results of these studies could be caused by different severity of diseases, the patient populations, the time periods for which the investigators included the patients [58]. Although the four cohort studies included in this study were of high quality, the results could not be combined due to the difference in their outcome indicators. The search in ClinicalTrials.gov showed that no study investigated the efficacy of IVIG as a therapeutic agent [59].

### 4.2. Strengths and limitations

This study is the first systematic review accessing the remdesivir, glucocorticoids and IVIG in treating children and adolescents with COVID-19. The study highlights the current status of evidence, identifies research gaps and proffer recommendations for developing clinical practice guidelines in treating children and adolescents with COVID-19. However, there are also some limitations: 1) All the studies using remdesivir in treating children with were low-quality single-arm cohort studies; thus, its efficacy and safety could not be clearly ascertained. 2) Due to the small sample size in the studies using glucocorticoids as treatment included in the study, the results of meta-analysis may be biased to some extent, and 3) Quantitative analysis of studies on the treatment of MIS-C by IVIG was not feasible due to the heterogeneity of their outcome indicators.

### 4.3. Further suggestions

Based on the results of this systematic review, we recommend 1) high-quality randomized controlled trials of potentially effective drugs for children with COVID-19; 2) develop better guidelines based on substantial current evidence, provide a timely guide for clinical workers, and update them in real-time according to the evidence situation.

## 5. Conclusion

Overall, the current evidence in the included studies is insignificant and of low quality, which does not adequately demonstrate the effectiveness and safety of using remdesivir, glucocorticoids, and IVIG in treating children and adolescents with COVID-19 or MIS-C. Therefore, it is recommended to conduct high-quality randomized controlled trials to provide substantial evidence for the development of guidelines.

## Data Availability

All data can be available referred to our manuscript.

## 6. Acknowledgments

## Authorship

All named authors meet the International Committee of Medical Journal Editors (ICMJE) criteria for authorship for this article, take responsibility for the integrity of the work as a whole, and have given their approval for this version to be published.

## Author contributions

Yaolong Chen, Enmei Liu, Zhengxiu Luo, Zijun Wang, Siya Zhao and Yuyi Tang contributed to the idea for the article. Zijun Wang, Siya Zhao, Yuyi, Zhili Wang, Qianling Shi, Lidan Gan, Shuai Peng and Weiguo Li performed the literature search data selection, data collection and study evaluation. Xiangyang Dang participated in the data analysis. Rafael González Cortés provided the data of one of the include study. Zijun Wang, Siya Zhao and Yuyi Tang drafted the manuscript. Yaolong Chen, Enmei Liu, Zhengxiu Luo, Qi Zhou, Qinyuan Li and Joy James Mafiana reviewed and provided feedback on the manuscript. All authors approved the final version of the manuscript. The corresponding author attests that all listed authors meet authorship criteria and that no others meeting the criteria have been omitted.

## Compliance with ethics guidelines

This article is based on previously conducted studies and does not contain any studies with human participants or animals performed by any of the authors.

## Data Availability

The datasets used and analyzed during this study are available from the corresponding author on reasonable request.

