## Supplementary File 3 for "Potentially effective drugs for the treatment of COVID-19 or MIS-C in children: a systematic review"

**Supplementary File 2 PRISMA 2020 flow diagram**

Clinical question 1 (remdesivir)

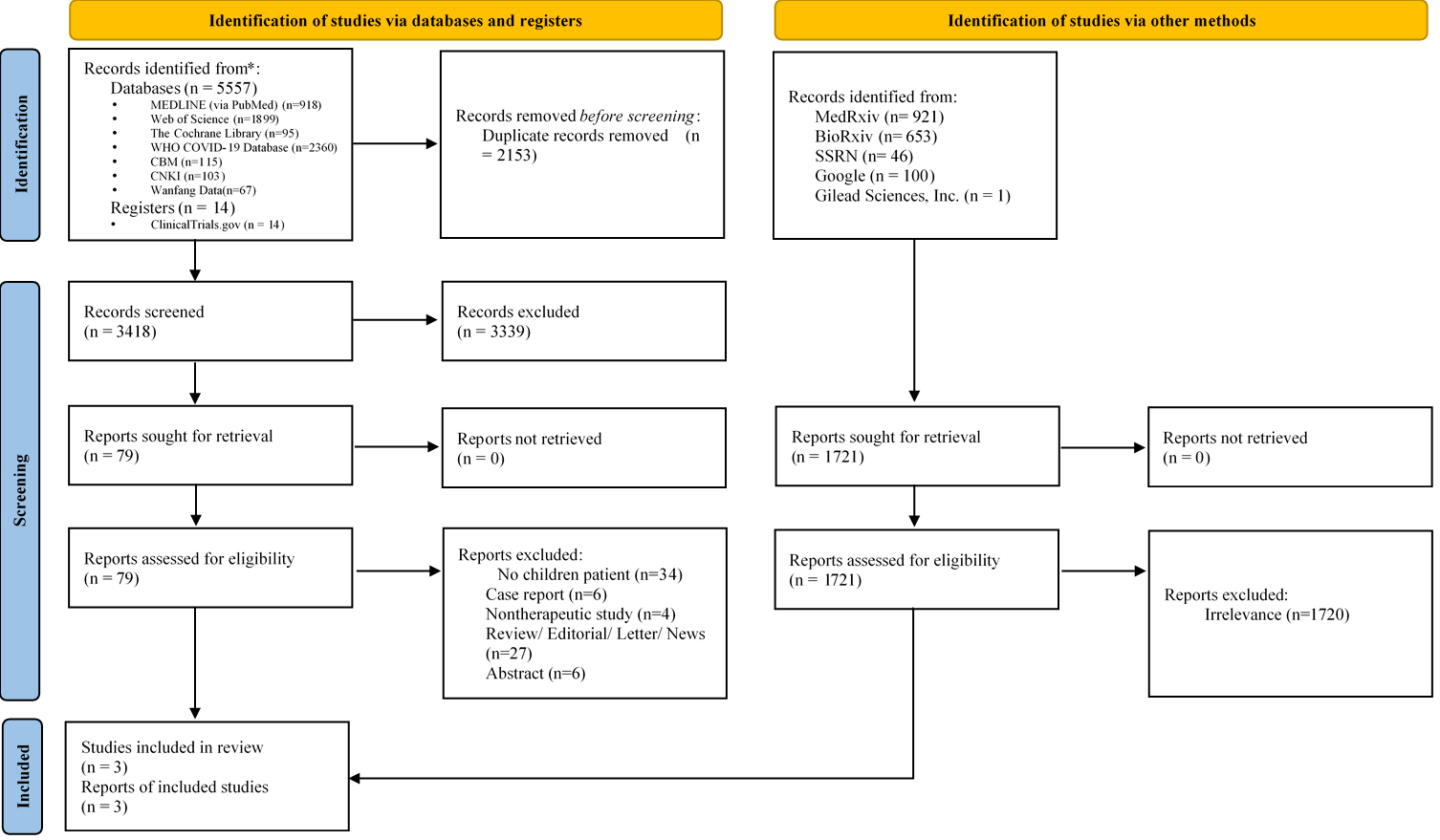

Clinical question 2 (glucocorticoids)

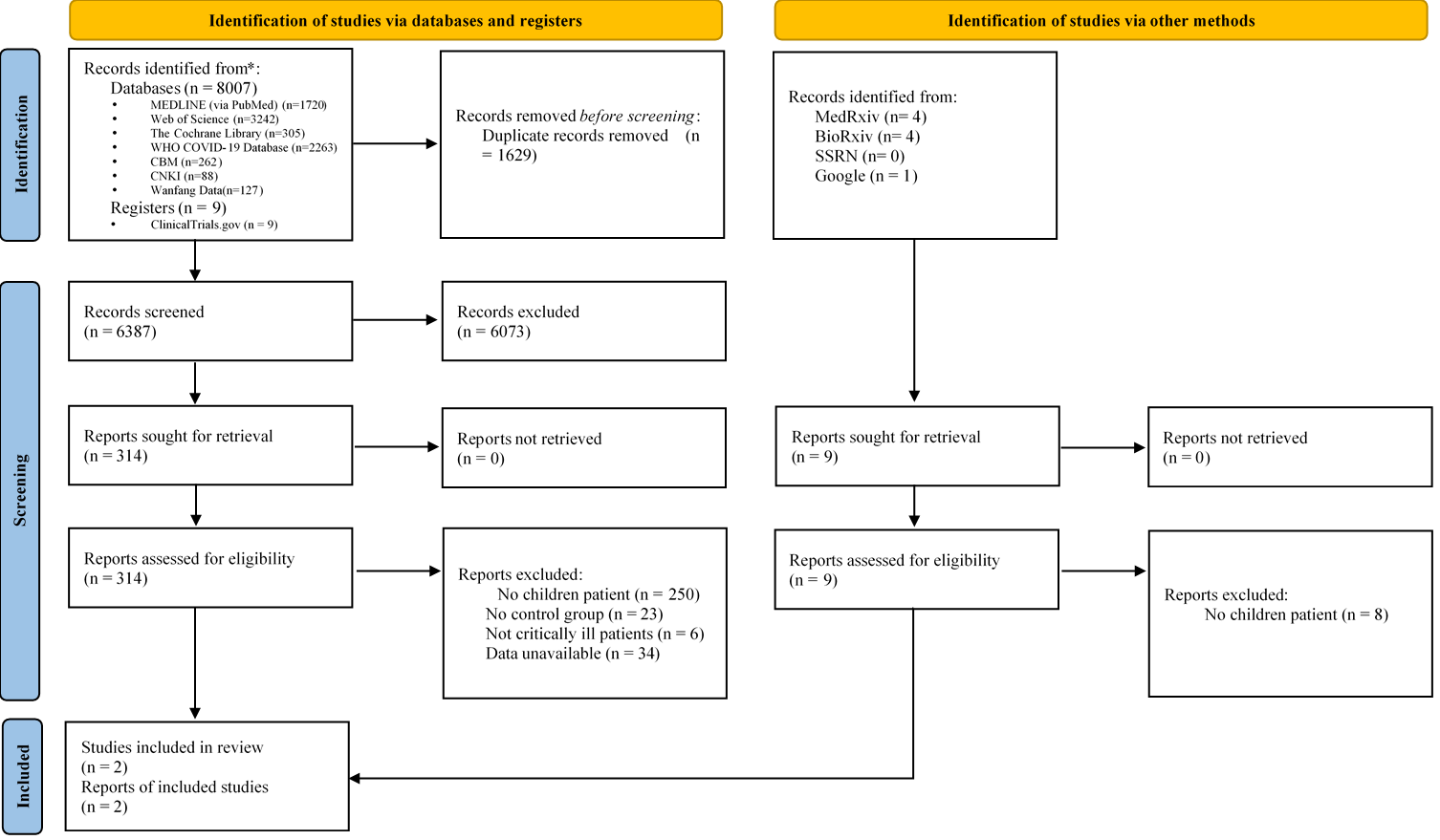

Clinical question 3 (IVIG)

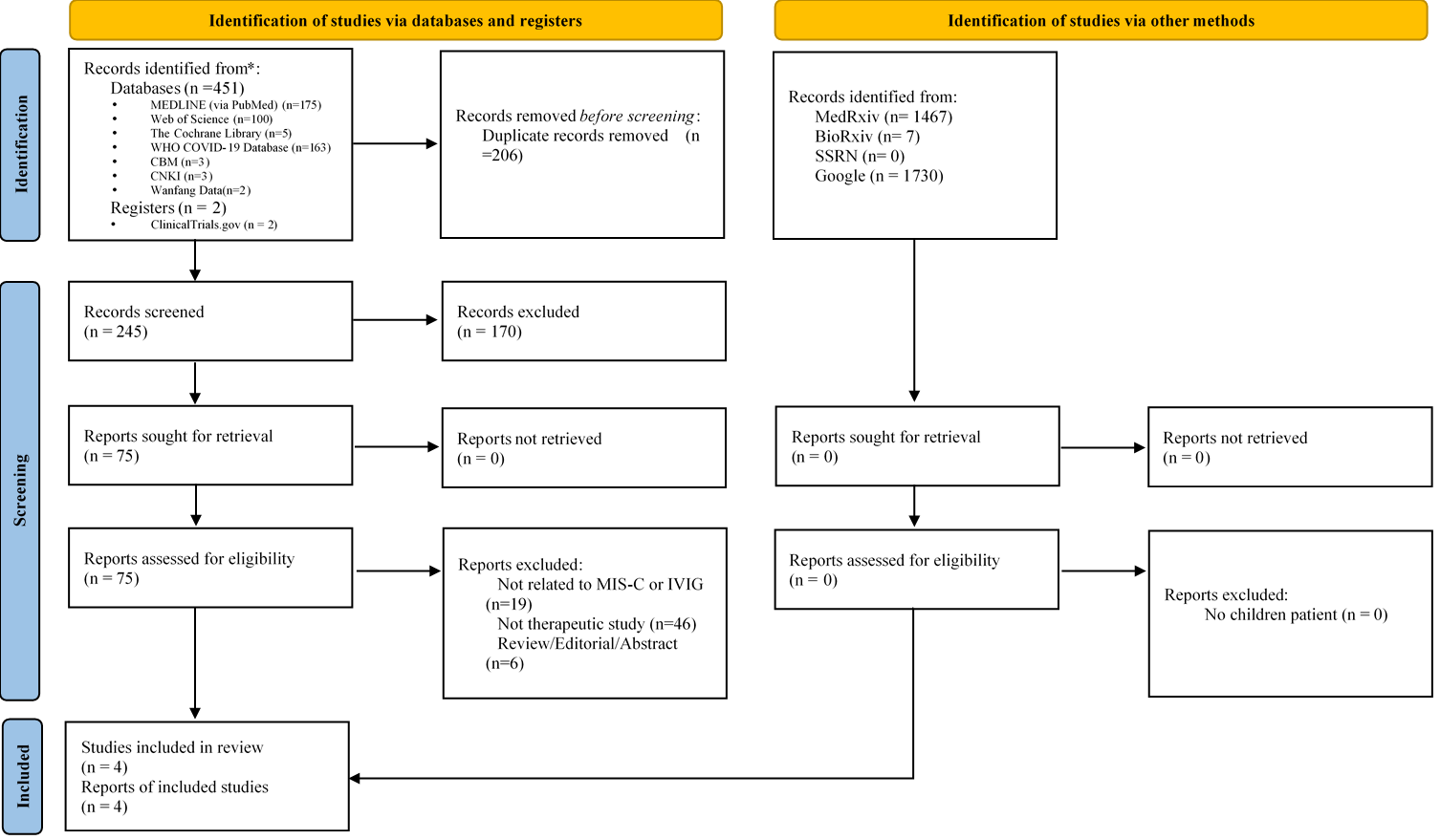
