## Supplementary File 4 for "Potentially effective drugs for the treatment of COVID-19 or MIS-C in children: a systematic review"

**Supplementary File 3** **Risk of Bias**

| **Table 1. Newcastle-Ottawa Scale** | | | | | | | | |
| --- | --- | --- | --- | --- | --- | --- | --- | --- |
| **Study ID** | **SELECTION** | | | | **COMPARABILITY** | **OUTCOME** | | |
|  | **Representativeness of the Exposed Cohort** | **Selection of the Non-Exposed Cohort** | **Ascertainment of Exposure** | **Demonstration That Outcome of Interest Was Not Present at Start of Study** | **Comparability of Cohorts on the Basis of the Design or Analysis** | **Assessment of Outcome** | **Was Follow-Up Long Enough for Outcomes to Occur** | **Adequacy of Follow Up of Cohorts** |
| Méndez-Echevarría et al. 2020 | * | / | / | * | / | / | * | * |
| Munoz et al. 2021 | * | / | * | * | * | / | * | * |
| Goldman et al. 2021 | * | / | * | * | * | * | * | * |
| García-Salido et al. 2020 | * | * | * | * | / | * | * | * |
| Ouldali et al. 2021 | * | * | * | * | * | * | * | * |
| Belhadjer et al. 2020 | * | * | * | * | / | * | / | * |
| Son et al. 2021 | * | * | * | * | / | * | / | * |
| McArdle et al. 2021 | * | * | * | * | / | * | * | * |
| * means one asterisk, which have been explained in the part of the “Risk of bias assessment” | | | | | | | | |

| **Table 2. Institute of Health Economics Scale** | | | | | | | | | | | | | | | | | | | | |
| --- | --- | --- | --- | --- | --- | --- | --- | --- | --- | --- | --- | --- | --- | --- | --- | --- | --- | --- | --- | --- |
| **Study ID** | **Domain 1** | **Domain 2** | | | **Domain 3** | | | **Domain 4** | | **Domain 5** | | | | **Domain 6** | **Domain 7** | | | | | **Domain 8** |
|  | **Item1** | **Item2** | **Item3** | **Item4** | **Item5** | **Item6** | **Item7** | **Item8** | **Item9** | **Item10** | **Item11** | **Item12** | **Item13** | **Item14** | **Item15** | **Item16** | **Item17** | **Item18** | **Item19** | **Item20** |
| Sun et al. 2020 | Yes | Unclear | No | Yes | Yes | Partial | Yes | Partial | Yes | Yes | Unclear | Yes | No | Yes | No | No | No | No | Yes | Yes |
| Domain 1: Study objective, including: 1. Was the hypothesis/aim/objective of the study clearly stated?; Domain 2: Study design, including: 2. Was the study conducted prospectively?; 3.Were the cases collected in more than one centre?; 4.Were patients recruited consecutively?; Domain 3: Study population, including: 5. Were the characteristics of the patients included in the study described?; 6.Were the eligibility criteria (i.e. inclusion and exclusion criteria) for entry into the study clearly stated?; 7.Did patients enter the study at a similar point in the disease?; Domain 4: Intervention and co-intervention, including: 8. Was the intervention of interest clearly described?; 9.Were additional interventions (co-interventions) clearly described?; Domain 5: Outcome measure, including: 10. Were relevant outcome measures established a priori?; 11.Were outcome assessors blinded to the intervention that patients received?; 12.Were the relevant outcomes measured using appropriate objective/subjective methods?; 13.Were the relevant outcome measures made before and after the intervention?; Domain 6: Statistical analysis, including: 14. Were the statistical tests used to assess the relevant outcomes appropriate?; Domain 7: Results and conclusions, including: 15. Was follow-up long enough for important events and outcomes to occur?; 16.Were losses to follow-up reported?; 17.Did the study provided estimates of random variability in the data analysis of relevant outcomes?; 18.Were the adverse events reported?; 19.Were the conclusions of the study supported by results?; Domain 8: Competing interests and sources of support, including: Were both competing interests and sources of support for the study reported? | | | | | | | | | | | | | | | | | | | | |
