## Supplementary File 5 for "Potentially effective drugs for the treatment of COVID-19 or MIS-C in children: a systematic review"

**Supplementary File 4 GRADE assessment (Summary of Findings table)**

| Table 1. Remdesivir | | | | | | | | | |
| --- | --- | --- | --- | --- | --- | --- | --- | --- | --- |
| № of studies | Certainty assessment | | | | | № of patients | | Effect Value  (95% CI) | Certainty |
|  | Risk of bias | Inconsistency | Indirectness | Imprecision | Other considerations | Total | Event |  |  |
| Adverse events | | | | | | | | | |
| Single-arm Cohort (3) | Serious^1^ | Not serious | Not serious | Serious^2^ | None | 112 | 46 | 37.1% (0.0%, 74.5%) | ⨁◯◯◯  VERY LOW |
| Serious adverse events | | | | | | | | | |
| Single-arm Cohort (3) | Serious^1^ | Not serious | Not serious | Serious^2^ | None | 112 | 21 | 16.2% (1.8%, 30.5%) | ⨁◯◯◯  VERY LOW |
| Extra-corporeal membrane oxygenation (ECMO) or invasive mechanical ventilation (IMV) | | | | | | | | | |
| Single-arm Cohort (3) | Serious^1^ | Not serious | Not serious | Serious^2^ | None | 112 | 45 | 37.2% (0%, 76.0%) | ⨁◯◯◯  VERY LOW |
| Mortality | | | | | | | | | |
| Single-arm Cohort (3) | Serious^1^ | Not serious | Not serious | Serious^2^ | None | 112 | 7 | 5.9% (1.5%, 10.2%) | ⨁◯◯◯  VERY LOW |

| Table 2-1. Glucocorticoids | | | | | | | | | | |
| --- | --- | --- | --- | --- | --- | --- | --- | --- | --- | --- |
| № of studies | Certainty assessment | | | | | № of patients | | | Effect Value  (95% CI) | Certainty |
|  | Risk of bias | Inconsistency | Indirectness | Imprecision | Other considerations | Sample | Intervention | Control |  |  |
| Mortality | | | | | | | | | | |
| Cohort (1)  Case series (1) | Serious^1^ | Not serious | Not serious | Serious^2^ | None | 69 | 2/40 | 0/21 | 2.79 (0.13, 60.87) | ⨁◯◯◯  VERY LOW |
| Mechanical ventilation rate | | | | | | | | | | |
| Cohort (1)  Case series (1) | Serious^1^ | Not serious | Not serious | Serious^2^ | None | 69 | 14/40 | 3/21 | 3.12 (0.80, 12.08) | ⨁◯◯◯  VERY LOW |

| Table 2-2. Glucocorticoids | | | | | | | | | | | | |
| --- | --- | --- | --- | --- | --- | --- | --- | --- | --- | --- | --- | --- |
| № of studies | Certainty assessment | | | | | № of patients | | | | | Effect Value  (95% CI) | Certainty |
|  | Risk of bias | Inconsistency | Indirectness | Imprecision | Other considerations | Sample | Intervention | | Control | |  |  |
|  |  |  |  |  |  |  | Mean | SD | Mean | SD |  |  |
| Duration of PICU admission | | | | | | | | | | | | |
| Cohort (1) | Serious^1^ | Not serious | Not serious | Serious^2^ | None | 69 | 6.9 | 8.2 | 4.9 | 3.5 | 2.0 (-0.95, 4.95) | ⨁◯◯◯  VERY LOW |

| Table 3-1. IVIG (IVIG + Glucocorticoids vs. IVIG) | | | | | | | | | | |
| --- | --- | --- | --- | --- | --- | --- | --- | --- | --- | --- |
| № of studies | Certainty assessment | | | | | № of patients | | | Effect Value  (95% CI) | Certainty |
|  | Risk of bias | Inconsistency | Indirectness | Imprecision | Other considerations | Sample | Intervention | Control |  |  |
| Treatment failure | | | | | | | | | | |
| Cohort (1) | Serious^1^ | Not serious | Serious^3^ | Not serious | None | 96 | 3/32 | 24/64 | 0.25 (0.09, 0.70) | ⨁◯◯◯  VERY LOW |
| Second-line treatment | | | | | | | | | | |
| Cohort (1) | Serious^1^ | Not serious | Serious^3^ | Not serious | None | 96 | 3/32 | 20/64 | 0.19 (0.06, 0.61) | ⨁◯◯◯  VERY LOW |
| Hemodynamic support | | | | | | | | | | |
| Cohort (1) | Serious^1^ | Not serious | Serious^3^ | Not serious | None | 96 | 2/32 | 15/64 | 0.21 (0.06, 0.76) | ⨁◯◯◯  VERY LOW |
| LVEF <55% | | | | | | | | | | |
| Cohort (1) | Serious^1^ | Not serious | Serious^3^ | Not serious | None | 52 | 2/12 | 14/40 | 0.20 (0.06, 0.66) | ⨁◯◯◯  VERY LOW |

| Table 3-2. IVIG (IVIG + Glucocorticoids vs. IVIG) | | | | | | | | | | | | |
| --- | --- | --- | --- | --- | --- | --- | --- | --- | --- | --- | --- | --- |
| № of studies | Certainty assessment | | | | | № of patients | | | | | Effect Value  (95% CI) | Certainty |
|  | Risk of bias | Inconsistency | Indirectness | Imprecision | Other considerations | Sample | Intervention | | Control | |  |  |
|  |  |  |  |  |  |  | Mean | SD | Mean | SD |  |  |
| Duration of PICU admission | | | | | | | | | | | | |
| Cohort (1) | Serious^1^ | Not serious | Serious^3^ | Not serious | None | 96 | 3.6 | 2.3 | 6.1 | 3.4 | -2.4 (-4.0, -0.7) | ⨁◯◯◯  VERY LOW |
| Time to recovery of left ventricle ejection fraction | | | | | | | | | | | | |
| Cohort (1) | Serious^1^ | Not serious | Serious^3^ | Not serious | None | 22 | 2.9 | NR | 5.4 | NR | NR | ⨁◯◯◯  VERY LOW |
| Isovolumic relaxation time | | | | | | | | | | | | |
| Cohort (1) | Serious^1^ | Not serious | Serious^3^ | Not serious | None | 22 | 6.4 | NR | 20.6 | NR | NR | ⨁◯◯◯  VERY LOW |
| Duration of PICU stay | | | | | | | | | | | | |
| Cohort (1) | Serious^1^ | Not serious | Serious^3^ | Not serious | None | 22 | 3.4 | NR | 5.3 | NR | NR | ⨁◯◯◯  VERY LOW |

| Table 3-3. IVIG (IVIG + Glucocorticoids vs. IVIG) | | | | | | | | | | |
| --- | --- | --- | --- | --- | --- | --- | --- | --- | --- | --- |
| № of studies | Certainty assessment | | | | | № of patients | | | Effect Value  (95% CI) | Certainty |
|  | Risk of bias | Inconsistency | Indirectness | Imprecision | Other considerations | Sample | Intervention | Control |  |  |
| Cardiovascular dysfunction on or after day 2 | | | | | | | | | | |
| Cohort (1) | Serious^1^ | Not serious | Serious^3^ | Not serious | None | 206 | 18/103 | 32/103 | 0.56 (0.34, 0.94) | ⨁◯◯◯  VERY LOW |
| Left ventricular dysfunction | | | | | | | | | | |
| Cohort (1) | Serious^1^ | Not serious | Serious^3^ | Not serious | None | 150 | 6/75 | 13/75 | 0.46 (0.19, 1.15) | ⨁◯◯◯  VERY LOW |
| Shock resulting in vasopressor use | | | | | | | | | | |
| Cohort (1) | Serious^1^ | Not serious | Serious^3^ | Not serious | None | 204 | 13/102 | 24/102 | 0.54 (0.29, 1.00) | ⨁◯◯◯  VERY LOW |
| Use of adjunctive therapy | | | | | | | | | | |
| Cohort (1) | Serious^1^ | Not serious | Serious^3^ | Not serious | None | 212 | 36/106 | 74/106 | 0.49 (0.36, 0.65) | ⨁◯◯◯  VERY LOW |

| Table 3-4. IVIG (IVIG + Glucocorticoids vs. IVIG) | | | | | | | | | | |
| --- | --- | --- | --- | --- | --- | --- | --- | --- | --- | --- |
| № of studies | Certainty assessment | | | | | № of patients | | | Effect Value  (95% CI) | Certainty |
|  | Risk of bias | Inconsistency | Indirectness | Imprecision | Other considerations | Sample | Intervention | Control |  |  |
| Receipt of inotropic support or mechanical ventilation on day 2 or later or death | | | | | | | | | | |
| Cohort (1) | Serious^1^ | Not serious | Serious^3^ | Not serious | None | 331 | 54/162 | 40/169 | 0.95 (0.37, 2.45) | ⨁◯◯◯  VERY LOW |
| Reduction in the score for disease severity on the ordinal scale by day 2 | | | | | | | | | | |
| Cohort (1) | Serious^1^ | Not serious | Serious^3^ | Not serious | None | 304 | 52/152 | 43/152 | 1.09 (0.53, 2.23) | ⨁◯◯◯  VERY LOW |

| Table 3-4. IVIG (Glucocorticoids vs. IVIG) | | | | | | | | | | |
| --- | --- | --- | --- | --- | --- | --- | --- | --- | --- | --- |
| № of studies | Certainty assessment | | | | | № of patients | | | Effect Value  (95% CI) | Certainty |
|  | Risk of bias | Inconsistency | Indirectness | Imprecision | Other considerations | Sample | Intervention | Control |  |  |
| Receipt of inotropic support or mechanical ventilation on day 2 or later or death | | | | | | | | | | |
| Cohort (1) | Serious^1^ | Not serious | Serious^4^ | Not serious | None | 237 | 12/68 | 40/169 | 0.30 (0.10, 0.85) | ⨁◯◯◯  VERY LOW |
| Reduction in the score for disease severity on the ordinal scale by day 2 | | | | | | | | | | |
| Cohort (1) | Serious^1^ | Not serious | Serious^4^ | Not serious | None | 212 | 16/60 | 43/152 | 1.95 (0.83, 4.60) | ⨁◯◯◯  VERY LOW |

Explanations

1. downgrade one level: The risk of bias is high due to the limitations of study design

2. downgrade one level: Sample size is less than optimal information sample (OIS) or confidence interval is too wide

3. downgrade one level: Glucocorticoids combined with IVIG

4. downgrade one level: Glucocorticoids vs. IVIG

CI: Confidence interval; NR: Not report;
